# Evidence suggests a novel cerebrospinal circulatory system exists in human nerves

**DOI:** 10.1101/2020.06.26.20141267

**Authors:** Joel E Pessa

**Affiliations:** 18 Florence Avenue, Arlington, Massachusetts 02476

## Abstract

Evidence from a 5-year study including 150 fresh human cadaver dissections, microsurgery, fluorescent microinjections, immunohistochemistry and confocal imaging suggests a novel CSF circulatory system exists in human nerves. We introduced this system in 2017. Here we provide further evidence to support our conclusion. No previous manuscript, text, or atlas has identified a CSF circulatory system in nerves. The human nervous system is devoid of lymphatics. CSF vessels in human nerves are distinct from blood vessels and lymphatics; may be a remnant of the primitive circulatory system in multi-cellular organisms; and likely predate the vascular circulation in animals.

## INTRODUCTION

Evidence from a 5-year study including 150 fresh human cadaver dissections, microsurgery, fluorescent microinjections, immunohistochemistry and confocal imaging suggests *a novel CSF circulatory system exists in human nerves*. We introduced this *system* in 2017.^1^ Here we provide further evidence to support our conclusion. No previous manuscript, text, or atlas has identified a CSF circulatory system in nerves.^2-22^ The human nervous system is devoid of lymphatics.^2-22^

This entire study began with the observation of vessels traveling on outer nerve (i.e. epineurial vessels) during carpal tunnel surgery (Figures 1a and b). These vessels had not been previously described.^2-22^ Our initial hypothesis was that epineurial vessels were part of the lymphatic system because they were devoid of red blood cells. This hypothesis turned out to be incorrect: we tried to characterize something “novel” in terms of what was already known (lymphatic vessels). In retrospect, this is probably the most important reason that the human CSF circulatory system has evaded detection until now: *it is a novel system comprised of novel vessels*. CSF vessels in humans likely predated the vascular circulation; they may be a remnant of the earliest circulatory system in multi-cellular organisms; and can be therefore characterized as *priangeo* (pri before; angeo vascular) *vessels*.

**Figure 1a.**
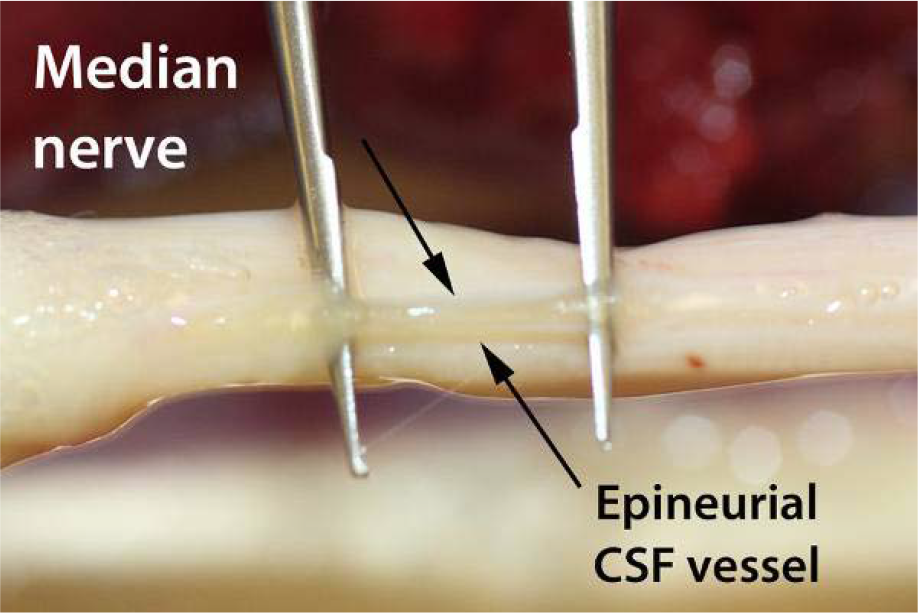
Epineurial CSF vessel on median nerve have not been previously described.

**Figure 1b.**
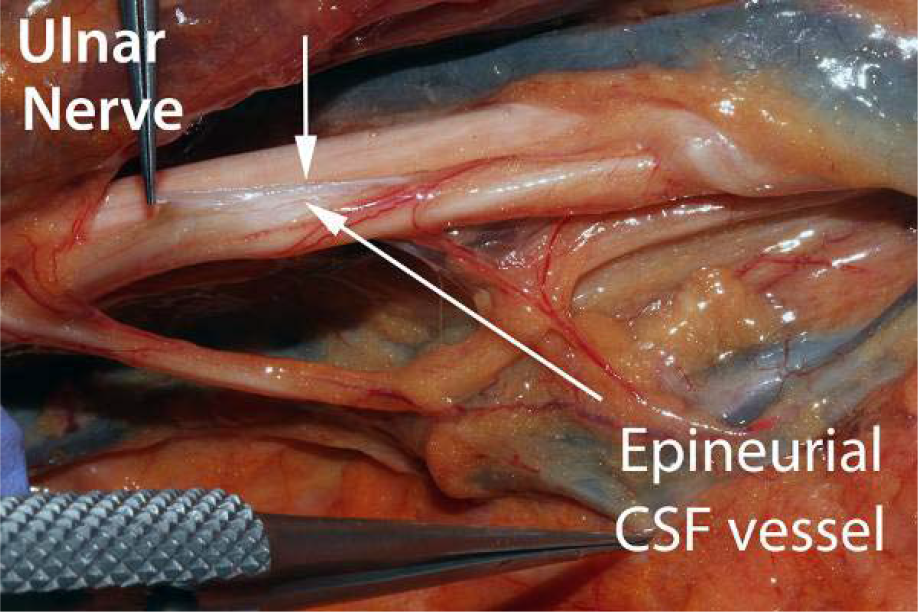
Epineurial CSF vessel on ulnar nerve above the elbow.

**Figure 1c.**
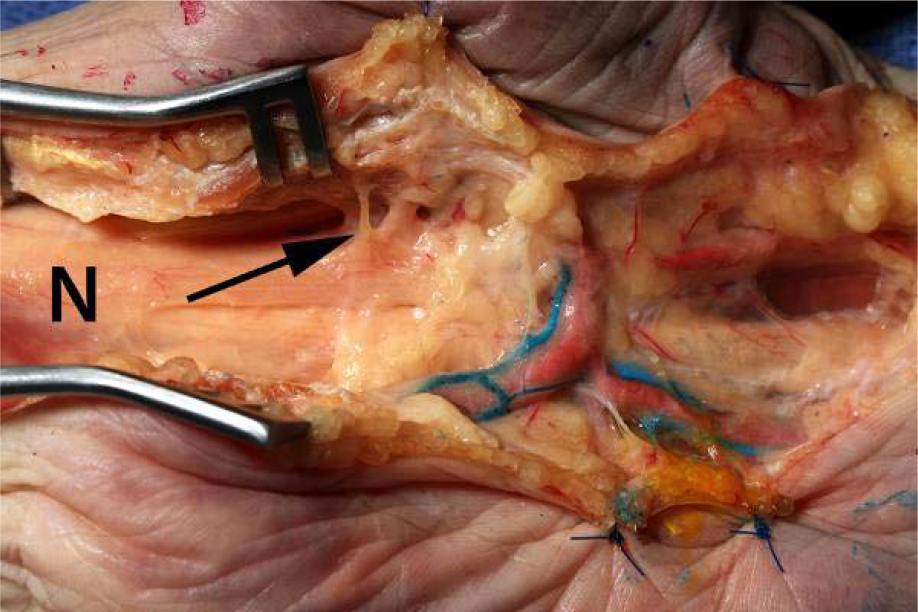
Epineurial CSF vessels drain to lymphatics via communicating branches (arrow) just proximal to potential sites of anatomic nerve compression. Median nerve (N) dissection in 67 year-old male specimen.

## RESULTS

### Ten studies were performed, each addressing a specific question

Techniques developed by Suami et al were adapted for this study.^23-26^ This manuscript uses terminology proposed by Sunderland.^20^ Nerves and brain are covered by layers of fascia: nerves by *neurium*; brain by *meninges* (singular meninx: membrane). Neurium and meninges are embryologically analogous layers. Note: peri*neural* means “surrounding the nerve”; peri*neurium* means “surrounding nerve fascicles”. The terms are not interchangeable.

### 1. Are epineurial CSF vessels present on all nerves?

**We identified epineurial CSF vessels in all 54 median/ulnar nerve dissections** (27 bilateral; 11f/16m; ages 58-98 years, mean 68.3). Epineurial CSF vessels travel along the entire length of nerves (Figures 1a and b). Epineurial CSF vessels communicate with adjacent lymphatics; the largest communications occur at sites of potential nerve compression (Figure 1c).

### 2. Do epineurial CSF vessels have a defined lumen?

**Dye injection confirmed that epineurial CSF vessels have a lumen in all 10 upper specimens tested** (6f/2m; ages 46 to 97 years; mean 76.6) (Figure 2). It was possible to inject epineurial CSF vessels with some macromolecular dyes (e.g. India ink), however, fluorescent dyes exhibited better flow.

**Figure 2.**
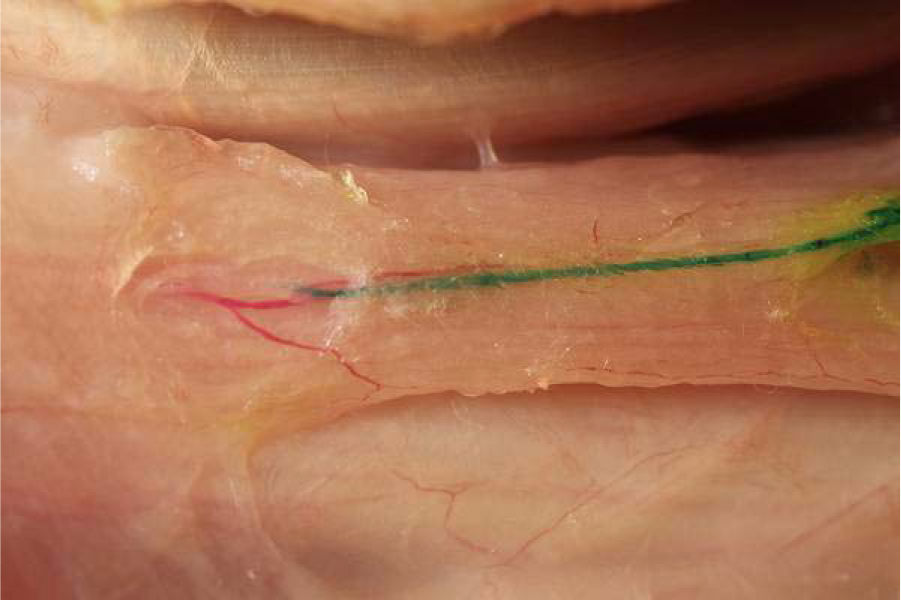
Injection of epineurial CSF vessel with green dye.

### 3. Do epineurial CSF vessels drain into thoracic duct?

**Vessels traveling from epineurial CSF vessels to the thoracic duct were identified in all 13 hemi-thoracic cadaver dissections** (5f/8m; ages 54-92; mean 74.3). Large communicating vessels were reliably found along the T1 nerve root (Figure 3a). FITC (fluoroscein isothiocyanate) injection in 2 specimens confirmed drainage into the thoracic duct. Epineurial CSF vessels on upper roots (C5, C6) first drain to the cervical CSF system and from there to thoracic duct (Figure 3b).

**Figure 3a.**
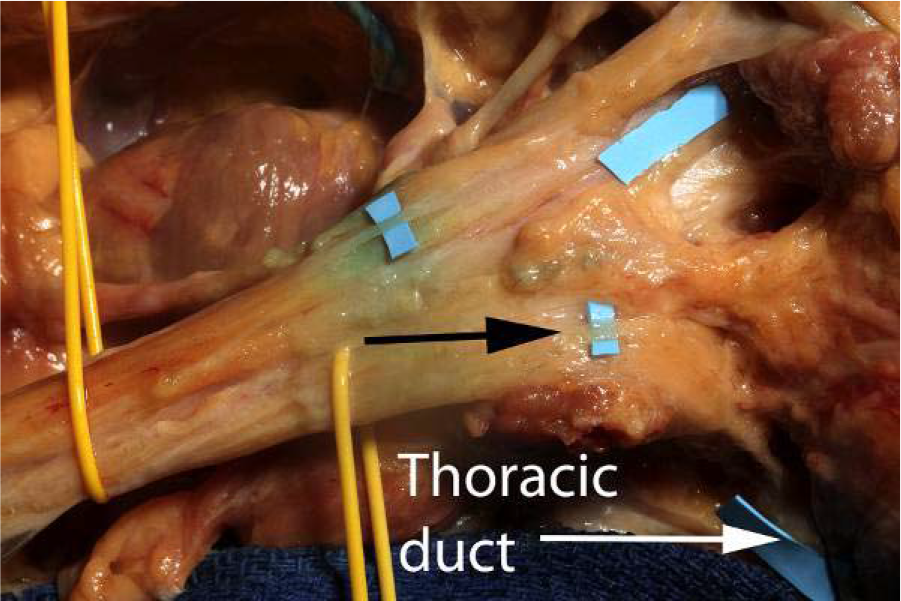
Brachial plexus dissection in 54 year-old male cadaver shows epineurial CSF vessels (black arrow) traveling to thoracic duct. FITC confirmed flow.

**Figure 3b.**
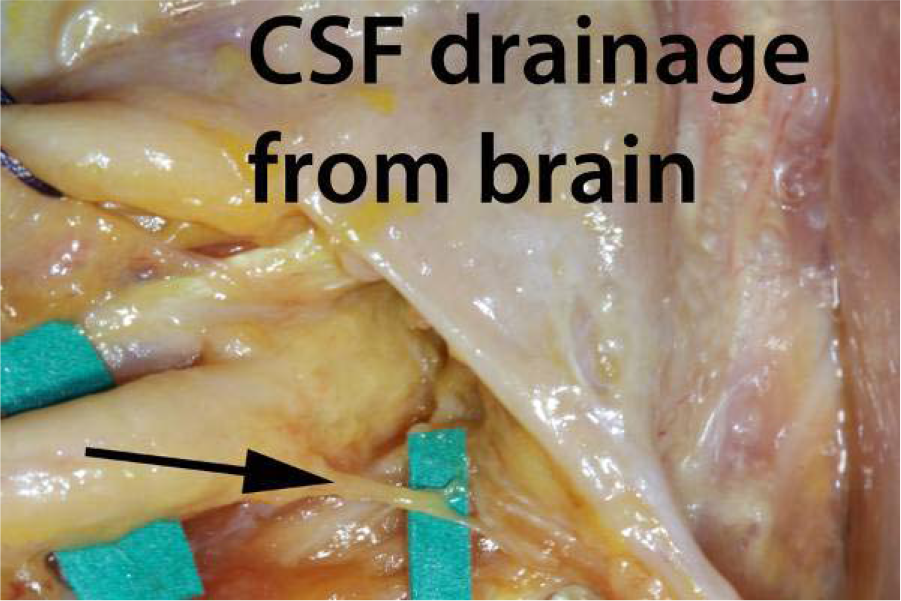
In upper root levels, epineurial CSF vessels (black arrow) traveled first to the cervical CSF drainage of the brain (within the carotid sheath) prior to draining into thoracic duct. 40 year-old female dissection is shown.

### 4. Do epineurial CSF vessels drain into lymphatics?

**Continuity between epineurial CSF vessels and the lymphatic system, and the superiority of fluorescent over visible dyes for imaging, was confirmed in all 8 upper extremity dissections** (6f/2m; ages 52 to 92 years. Mean 83). Raters were unable to document dye traveling from lymphatic to nerve using ambient light (0% of responses) (Figure 4a). Raters positively identified fluorescent dye traveling from lymphatics to epineurial CSF vessels in 75% of responses (Figure 4b). Fisher’s Exact Probability was significant with p < .0001. Inter-observer agreement was good between raters 1 and 2, and perfect (Kappa = 1) for raters 2 and 3 (see APPENDIX A).

**Figure 4a.**
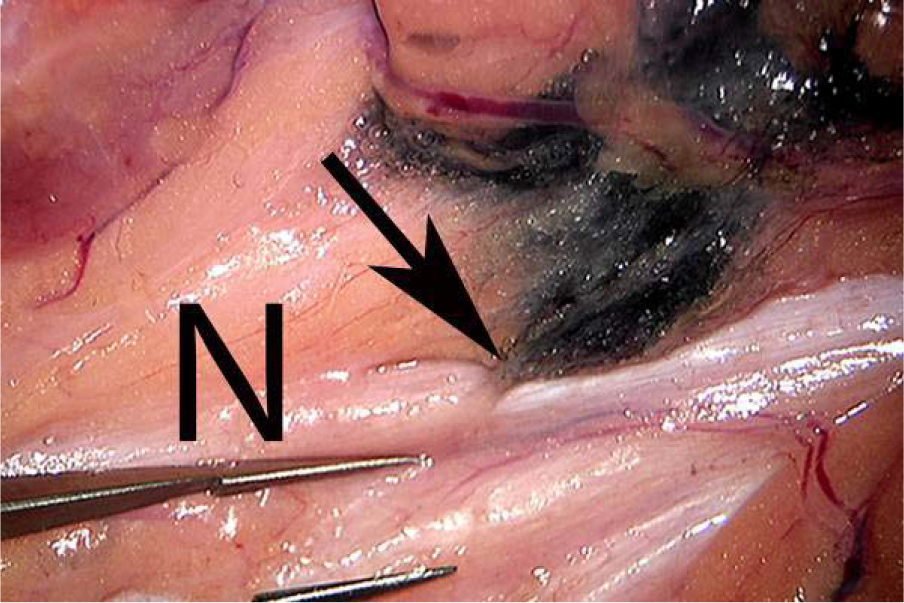
Raters were unable to identify dye injected into a lymphatic traveling to epineurial CSF vessels on median nerve using ambient light in this 73 year-old female dissection.

**Figure 4b.**
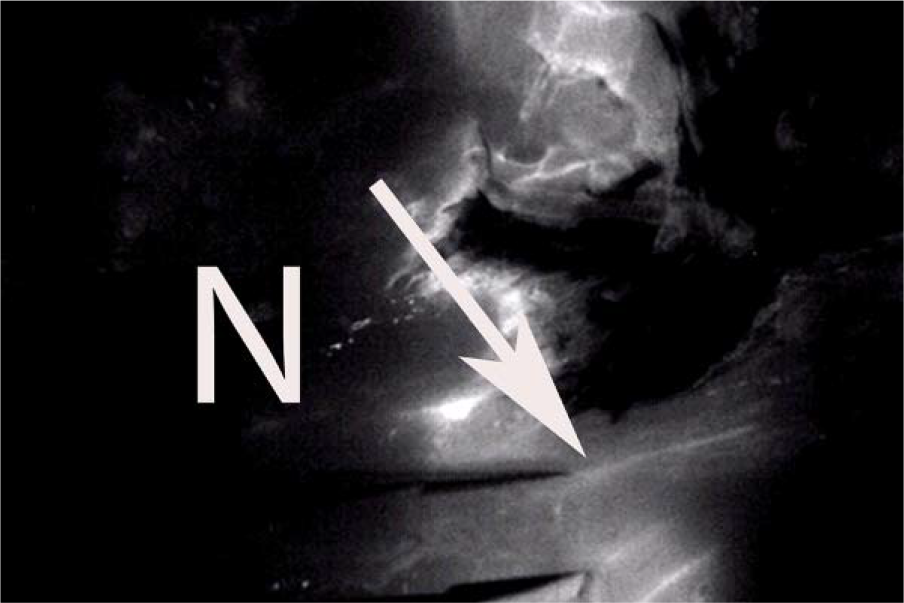
Raters identified dye traveling from lymphatic to epineurial vessels (white arrow) used infra-red imaging in 75% of their responses.

### 5. Do epineurial CSF vessels express a biomarker?

**Epineurial CSF vessels in all 28 median nerve specimens expressed vimentin** (14f/14 male; ages 52 to 95 years; mean 72.3). A representative section is shown (Figure 5).

**Figure 5.**
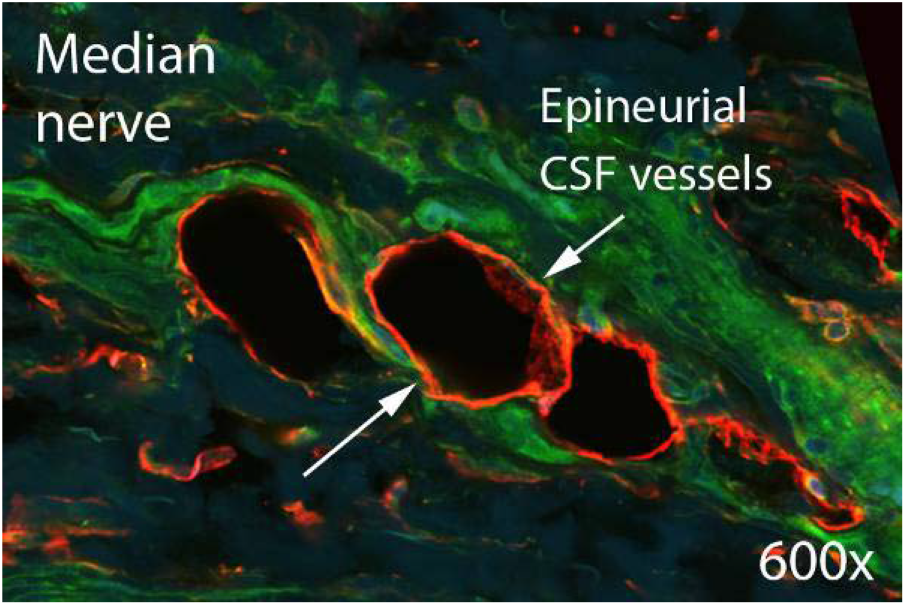
Epineurial CSF channels in median nerve from a 29 year-old female cadaver dissection are vimentin+.

### 6. Is vimentin a reliable biomarker for perineurial CSF vessels?

**Raters’ ability to identify perineurial fluorescence was reliable using vimentin** compared to either LYVE-1 or podoplanin using 8 median nerve specimens (2f/6m; ages 29 to 83 years; mean 75.3). The F-test suggested the 3 groups were significantly different (f=30.7; df1=2; df2=14; p < .0001; APPENDIX A). For added confirmation, perineurial fluorescence was compared using vimentin vs. LYVE-1 by averaged triplicate images of a region of interest (ROI). Vimentin was statistically superior to LYVE-1 (p < .0001, see APPENDIX A). Figure 6 shows a representative nerve section.

**Figure 6.**
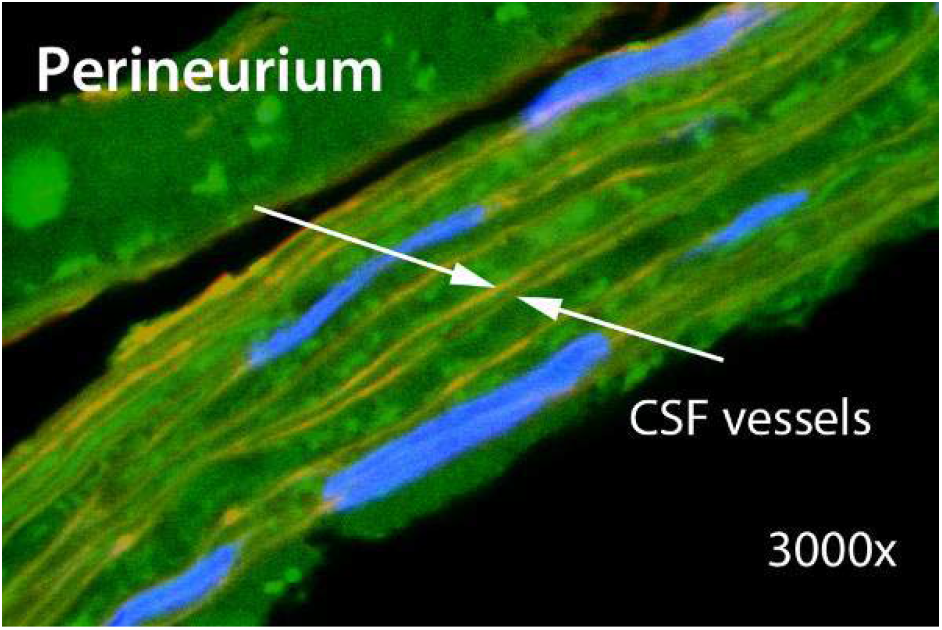
Perineurial CSF vessels are seen in median nerve at 3000x (Work from 2014).

### 7. Can we identify the CSF circulation of human brain by anatomic dissection?

**Both dural and arachnoid CSF vessels were identified in 13 cranial dissections** (8f/5m; ages 58 to 93 years; mean 72). Dural and arachnoid CSF vessels are clearly visible (Figure 7a). Dural CSF vessels may exit the skull through foramina (Figure 7b) and communicate with lymphatics at sites of periosteal fixation such as the nuchal ridge (Figure 7c). The main CSF channel in the sagittal sinus is identified (Figure 7d).

**Figure 7a.**
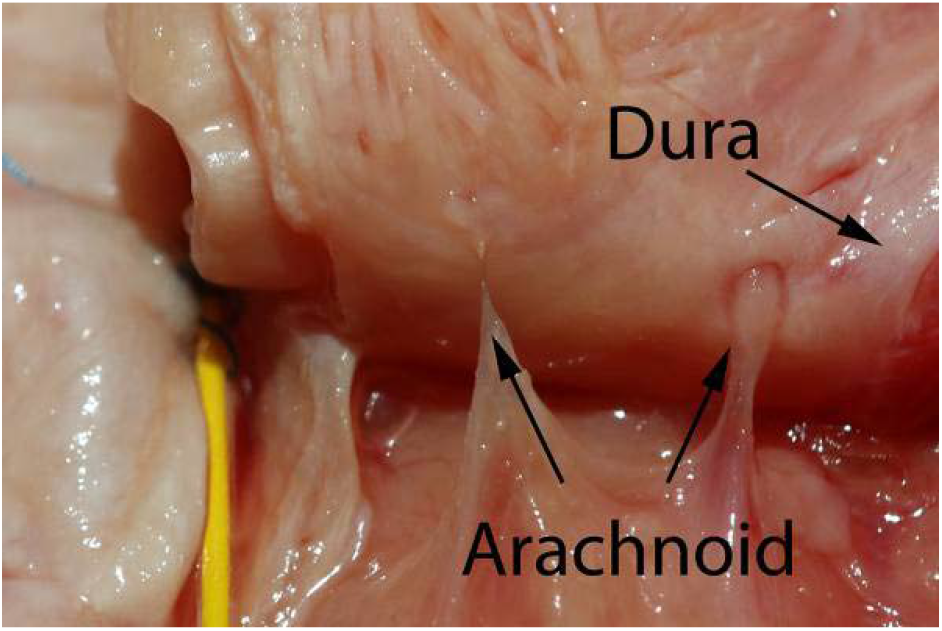
In this 92 y/o female cranial dissection, CSF vessels can be seen on dura (arrow). Arachnoid CSF vessels (arrow) travel to the dural CSF system with veins, a design that protects from shear.

**Figure 7b.**
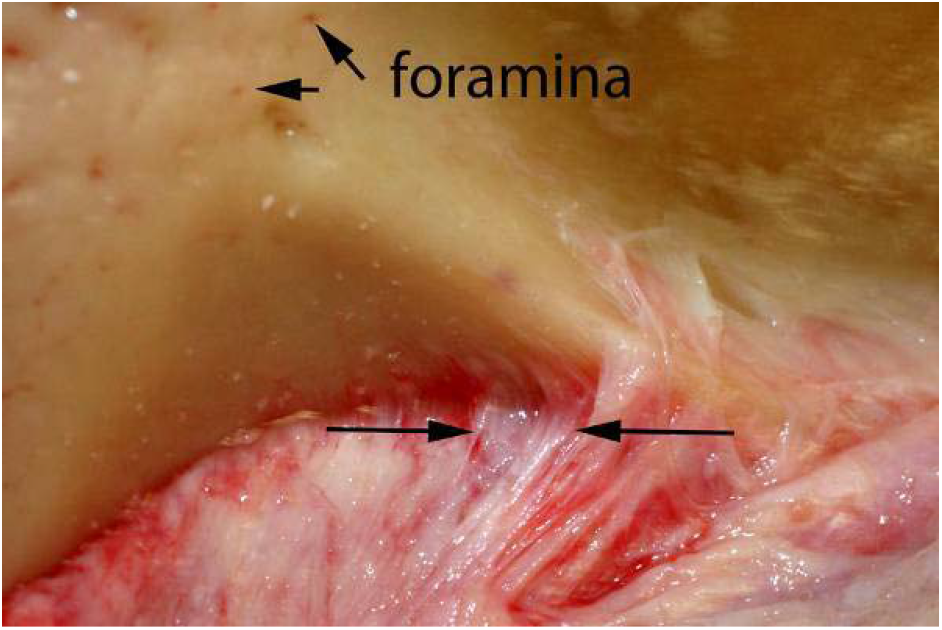
Dural CSF vessels (arrows) travel through foramina in the skull, seen in the upper left of this image of this 74 year-old female dissection.

**Figure 7c.**
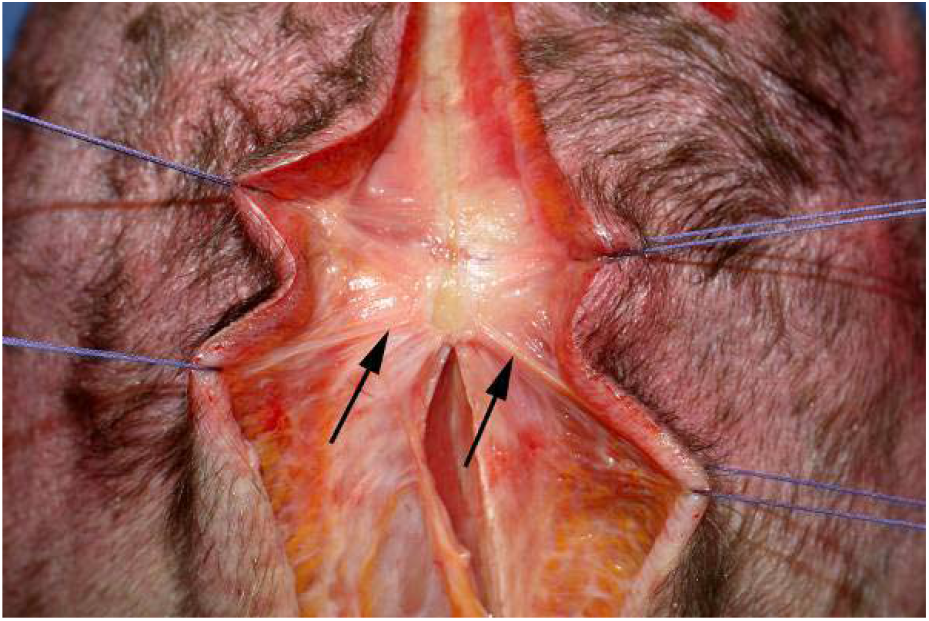
Dural CSF vessels exit the skull at sites of periosteal fixation where they drain into lymphatics (arrows), as seen here at the posterior nuchal ridge.

**Figure 7d.**
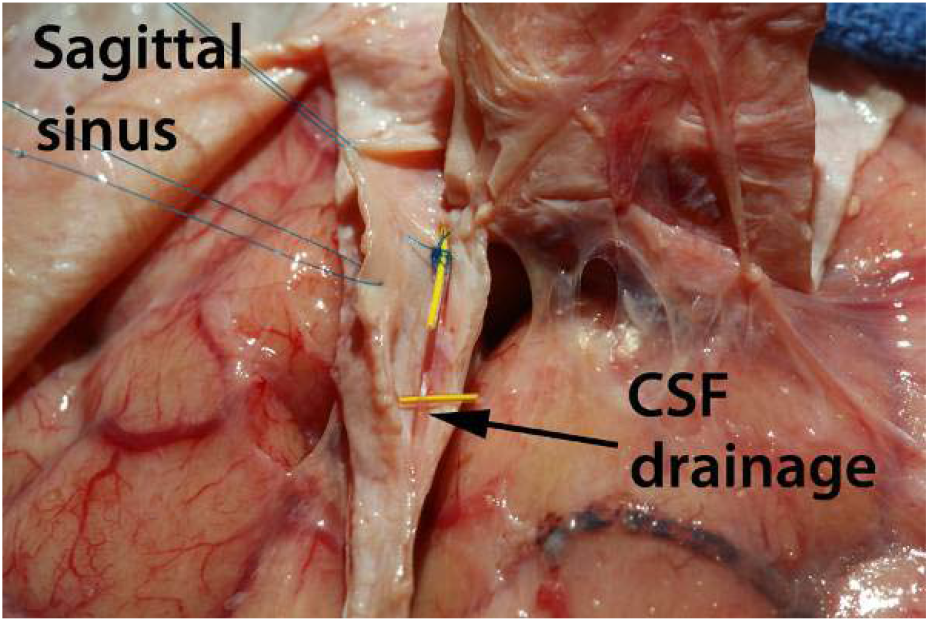
The main CSF channel in the (right hemisphere) traveling in sagittal sinus.

### 8. What is the terminal CSF drainage from the human brain?

**We identified terminal CSF draining vessels within the carotid sheath in all 13 anatomic dissections** (8f/5m; ages 35 to 87 years; mean 58.7). Cervical CSF vessels exit the skull through the CSF foramen, and travel along posterior internal jugular vein to the thoracic duct (Figure 8a). Osteotomies enabled us to dissect an enbloc specimen of the intra-cranial CSF system (from temporal fossa) in continuity with the cervical drainage system (Figure 8b).

**Figure 8a.**
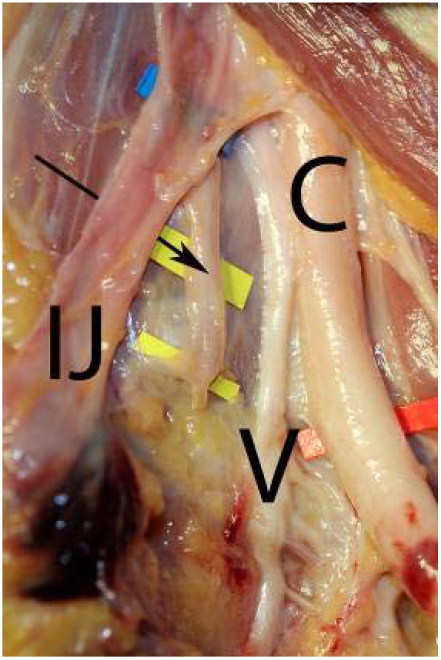
The entire cerebral CSF circulation drains into a plexus of vessels (arrow) traveling along the posterior internal jugular vein (IJ). Carotid artery (C) and vagus nerve (V) are noted.

**Figure 8b.**
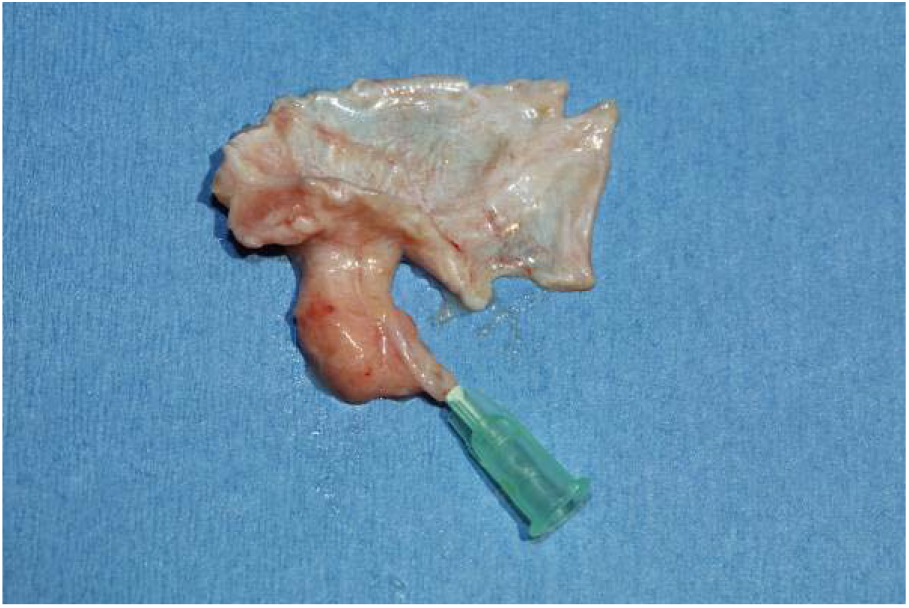
Cervical CSF drainage (cannulated) is shown with intra-cranial CSF system.

### 9. Does CSF drain from the brain to thoracic duct?

**Continuity between the intra-cranial and the extra-cranial CSF circulation and the superiority of fluorescent dye for visualization was documented in all 8 cadaver preparations** (7f/1m; ages 40 to 95 years; mean 87.6). Three raters were unable to identify visible dye traveling from the neck to the temporal fossa using visible dye and ambient light (Figure 9a). The same raters positively identified fluorescent dye after cervical injection traveling to the temporal fossa in 87.5% of their responses (Figure 9b). This was statistically significant with p < .0001 (Kappa = 1 for all observers).

**Figure 9a.**
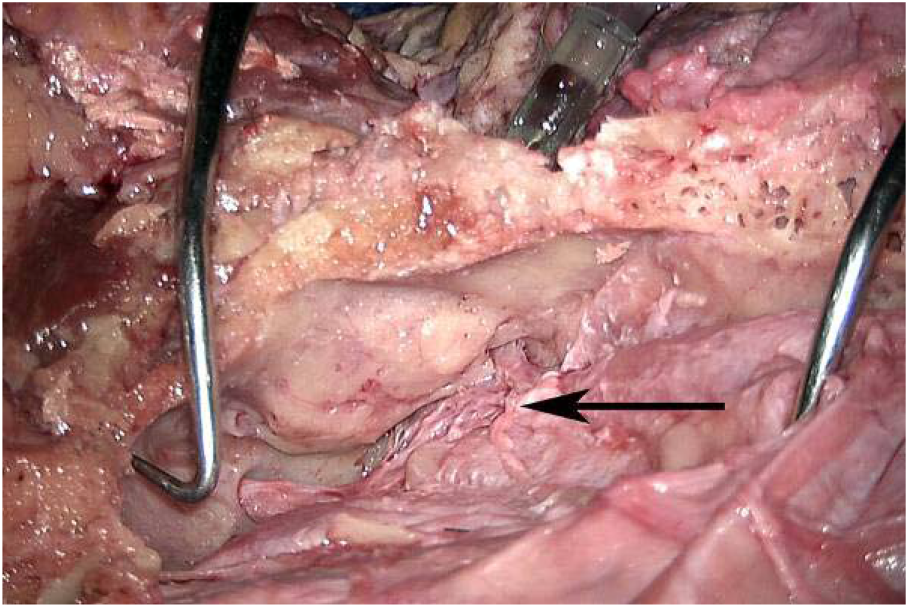
Raters were unable to visualize dye injected from the neck traveling to the terminal intra-cranial CSF circulation (arrow) using ambient light.

**Figure 9b.**
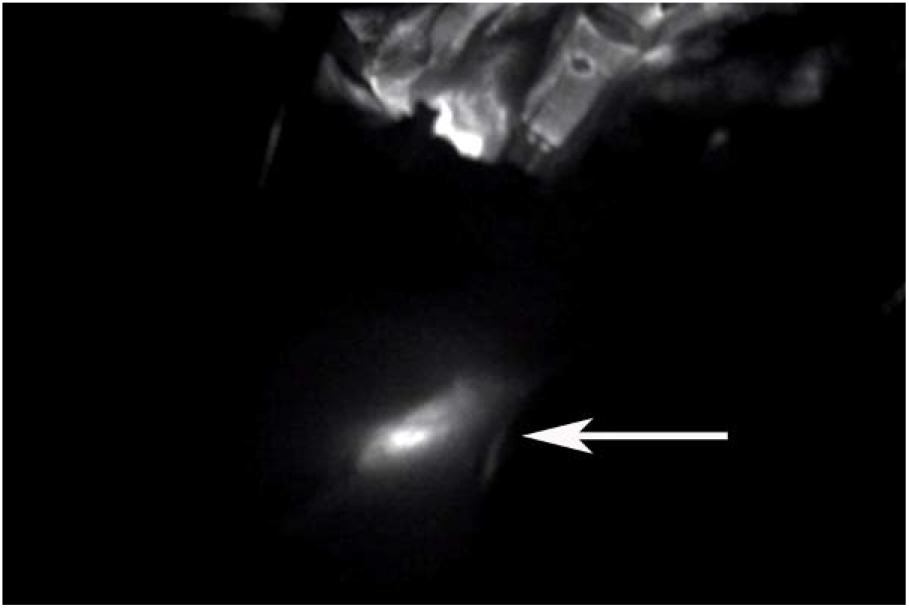
Raters identified dye traveling traveling retrograde from the neck to the terminal intra-cranial CSF system (arrow) in 87.5% of their responses. Note the syringe visible in upper photo containing IR800 dye.

### 10. What additional biomarkers are expressed by the CSF circulatory system?

**CSF vessels in human nerve express F-actin; GLUT-1; Claudin 1/3; and N-cadherin**. GLUT-1 expression was noted in endocortical CSF vessels, arachnoid CSF vessels, dura, and perineurial CSF vessels. This could be the structural correlate to the physiological blood-brain barrier (Figure 10).

**Figure 10.**
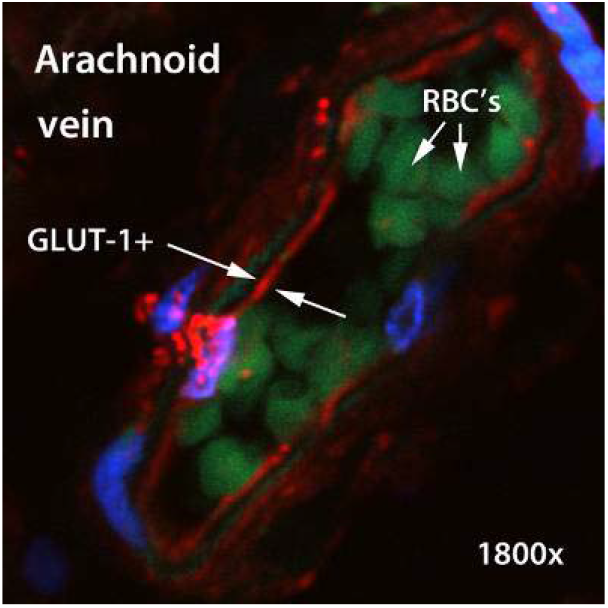
GLUT-1+ CSF paravascular channels in the wall of arachnoid vein are found in the human spinal cord. These CSF vessels represent the possible *structural* blood-brain barrier.

**Results are summarized in Table 1**.

**Table 1.** Studies from a series of 150 fresh cadaver dissections performed to address specific questions regarding the CSF circulatory system of human nerves (and brain).

| Question to be studied | #/Gender | Ages | Technique | Findings or conclusions |
| --- | --- | --- | --- | --- |
| Are epineurial CSF vessels present on all nerves? | 11f/16m<br>(N=54) | 58-98<br>(68.3) | Microsurgical dissection | Epineurial CSF vessels are present on all nerves |
| Do epineurial CSF vessels have a defined lumen? | 6f/2m<br>(N=8) | 46-97<br>(76.6) | Direct injection with dyes | Epineurial CSF vessels have a defined lumen |
| Do epineurial CSF vessels drain into thoracic duct? | 5f/8m<br>(N=13) | 54-92<br>(74.3) | Dissection; injection | Epineurial CSF vessels drain into thoracic duct |
| Does continuity exist with the lymphatic system? | 6f/2m<br>(N=8) | 52-92<br>(83.3) | Comparison of IR imaging | Continuity exists; infra-red imaging superior p<.0001 |
| Do epineurial CSF vessels express a biomarker? | 14f/14m<br>(N=28) | 52-95<br>(72.3) | Immuno-histochemistry | Epineurial CSF vessels express vimentin |
| Is vimentin a reliable biomarker for perineurial CSF vessels? | 2f/6m<br>(N=8) | 29-83<br>(75.3) | Immuno-histochemistry | Vimentin superior to LYVE-1 or podoplanin p<.0001 |
| Can the cerebral CSF circulation be visualized macroscopically? | 8f/5m<br>(N=13) | 58-93<br>(72.0) | Gross anatomy | Dural and arachnoid CSF vessels observed |
| Does the cerebral CSF circulation drain into the thoracic duct? | 8f/5m<br>(N=13) | 35-87<br>(58.7) | Retrograde dissection | Anatomical continuity from Thoracic duct to cranial base |
| Does continuity exist between the intra and extra cranial CSF circulation | 7f/1m<br>(N=8) | 40-95<br>(87.6) | Comparison Of IR imaging | Continuity exists; infra-red Imaging superior p<.0001 |
| What additional biomarkers are expressed by the CSF circulation? | Random sampling | 28-98 | Immuno-histochemistry | CSF vessels express F-actin, GLUT-1, Claudin 1/3, cadherin |

**Table 2.**
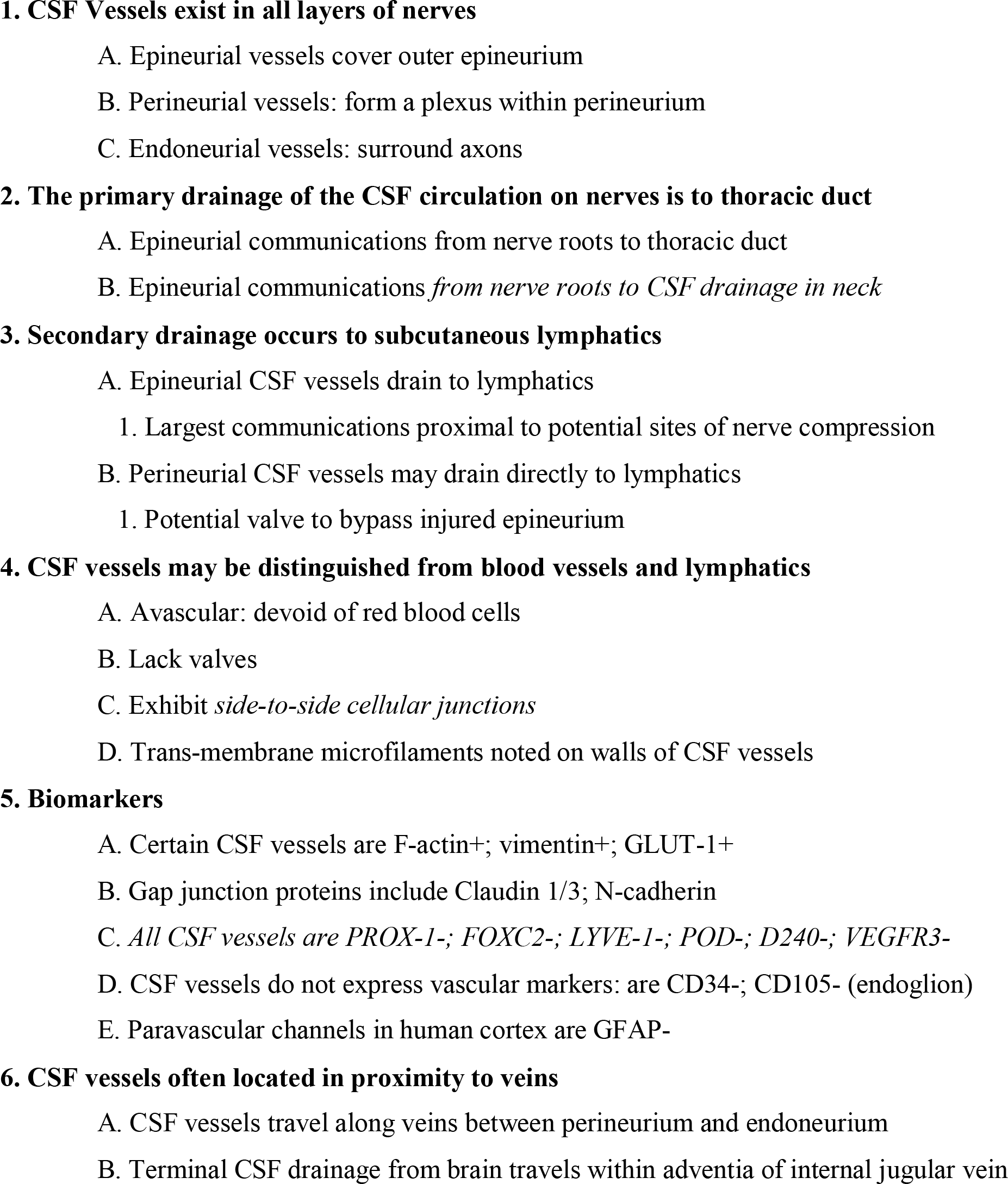
Features of the CSF circulatory system of human nerves.

**Table 3.**
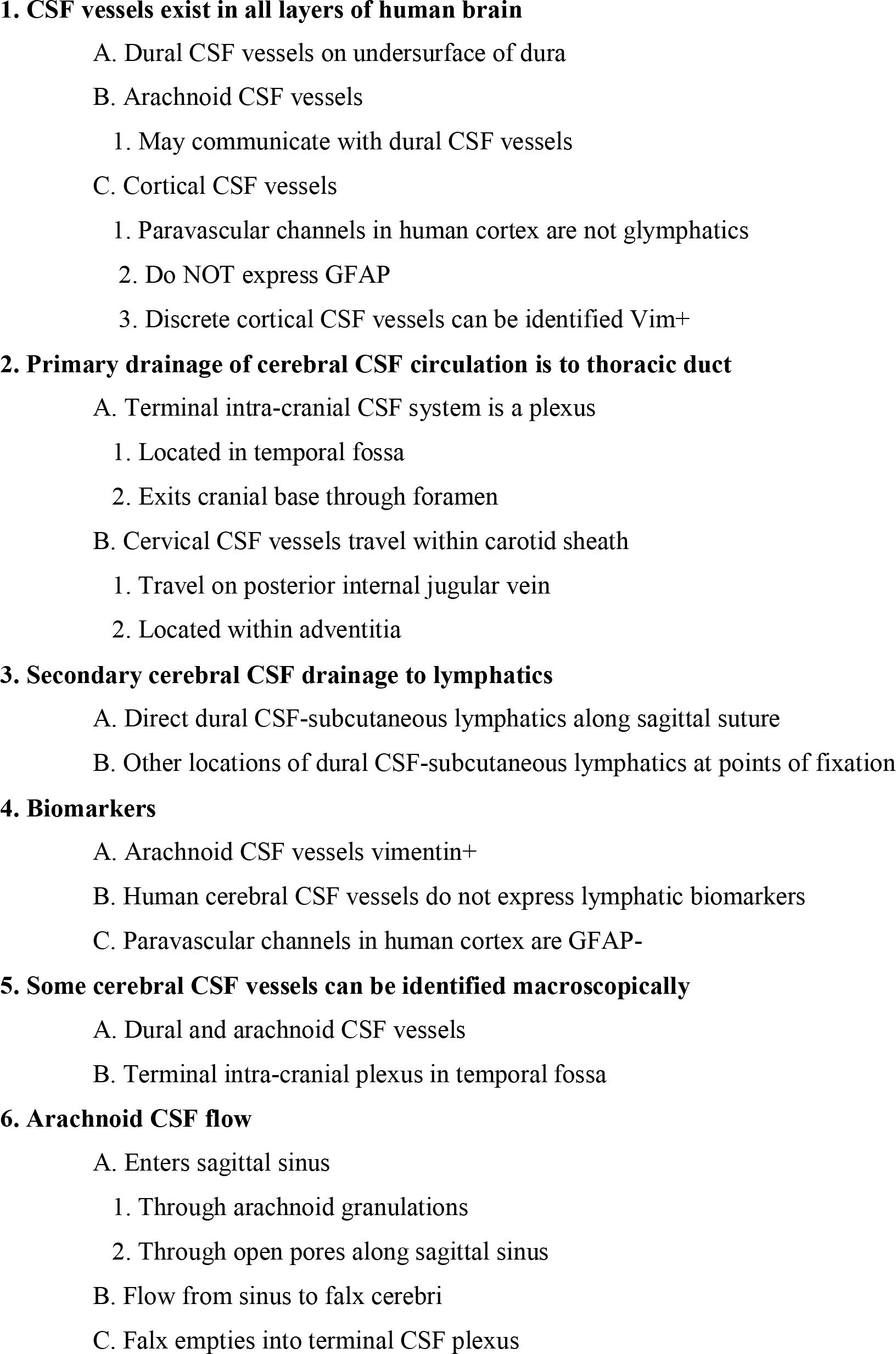
Features of the human cerebral CSF circulatory system.

## DISCUSSION

Evidence suggests a novel CSF circulatory system exists in all human nerves. To our knowledge no prior manuscript, textbook, or atlas has identified any vessels on either peripheral or central nerves with the exception of arteries and veins.^2-22^ The human nervous system is devoid of lymphatics.^2-22^

Characteristics of the CSF circulatory system on nerves include the following (Table 1):

1. CSF vessels exist in all layers of nerve (and brain);
2. Primary CSF drainage in humans is to thoracic duct;
3. Secondary CSF drainage exists to lymphatics;
4. CSF vessels can be distinguished from blood vessels/lymphatics; and
5. Biomarkers may include vimentin, GLUT-1, and F-actin.

### CSF vessels are distinct from blood vessels and lymphatics

CSF vessels *lack valves*, unlike blood vessels and lymphatics. The CSF circulatory system is *redundant* on several levels, a design that helps regulate pressure and flow. Primary drainage is to thoracic duct; secondary drainage to lymphatics. In addition, perineurial CSF vessels can bypass epineurium to drain into lymphatics. The same bypass is found in human brain and in spinal cord (Figure 11).

**Figure 11.**
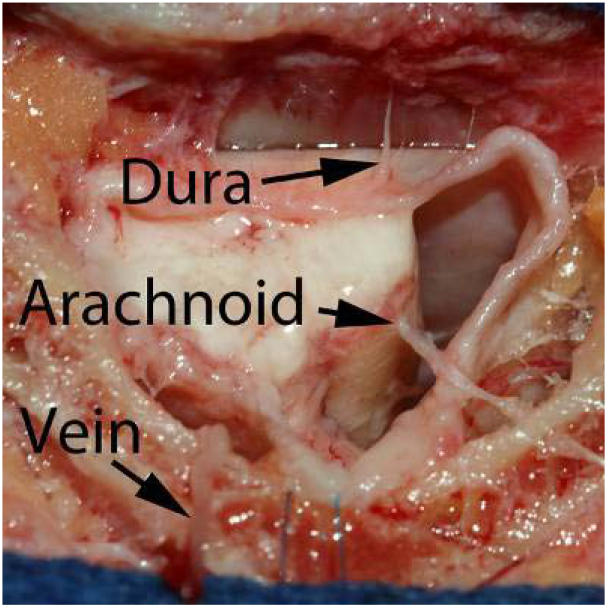
Redundancy of the human CSF circulatory system exists on several levels. Dural CSF vessels (arrow from dura) drain to subcutaneous lymphatics. Arachnoid CSF vessels can bypass dura (arrow from arachnoid) to drain directly into subcutaneous lymphatic vessels. 80 year-old male spinal cord dissection.

CSF vessels do not express any known lymphatic or vascular biomarkers.^27-31^ This is in contrast to Loveau et al’s work that characterized CSF vessels in *mouse dura* as lymphatics.^32^ Inter-species differences may exist. Human CSF vessels do not express glial fibrillary acidic protein (GFAP) in any layer of nerve or brain, and can not be characterized as glymphatic vessels found in mouse cortex by Iliff et al.^33^ Human CSF vessels express vimentin, a type-3 neurofilament protein usually expressed by cells of mesenchymal lineage.^34^ Another study suggests human perineurium expresses F-actin, vimentin, and GLUT-1, but those investigators were unable to identify CSF vessels.^35^ Much of the CSF circulatory system in the human brain can be visualized without magnification. Cerebrospinal fluid in the brain drains from sinus → falx → temporal fossa. The major CSF vessel on either side of the sagittal sinus is easily identified (Figure 7a). CSF ultimately drains into a plexus of vessels located within the temporal fossa (Figure 9a). The authors initially used loupes, micro-instruments, and the operating microscope to facilitate dissection. So it was a surprise (and humbling) to learn that *much of the human CSF circulatory system had been identified over 200 years ago*.

### A case of lost and found

We did our historical review at Harvard’s Countway Medical Library (Historical Medicine Section) and reviewed atlases by Cruickshank, Mascagni, and Poirier.^2-4, 7^ Mascagni writes as follows (1787):

> *Ut notrae vasorum lymphaticorum ichnographiae quod licuit, complementum accederet in postrema hac Tabula, quod in dura & pia meninge, & arachnoide, in cerebri, & cranii basi nobis occurrit… Occasione verò, qua alia figura vasorum cerebri, & meningum truncos juxta vasa sanguinea dexteri lateris e cranio prodeuntes, ac per collum excurrentes demonstrantur…in ultima icone totius sysematis terminus in venarum subclviae & internae utriusque lateris concursum re praefentatur*.

This translates as follows:

“(That) our illustration of the lymphatic vessels nears completion in the final Table, and that (the lymphatic vessels) in dura, pia, and arachnoid meninges of the brain meet in the cranial base…However occasionally (as seen in) the other figure of cerebral (blood) vessels *these meningeal vessels surround the right lateral vein* (sic) and exit from the brain to demonstrably travel along the neck. This entire system ultimately drains into the (confluence) of the subclavian and internal jugular veins”.

The vessels described by Mascagni (and by Cruickshank and Poirier) are the CSF circulatory system of the human brain.^2-4, 7^ Mascagni identified much of the CSF circulatory system in the human brain:

1. CSF vessels are present in all meningeal layers;
2. CSF vessels travel on the walls of blood vessels;
3. CSF vessels form a plexus in the temporal fossa; and
4. The entire CSF circulation of the human brain drains into thoracic duct.^3,4^

Mascagni’s only error was to characterize CSF vessels as lymphatics, most likely because their terminal drainage is thoracic duct (the known terminal drainage of all lymphatic vessels since Eustachius’ initial description in 1533). Mascagni, like many other investigators including ourselves, tried to characterize these novel vessels in terms of what was already known.

### Clinical implications

Evidence that a novel CSF circulatory system exist in human nerves may lead to improved understanding of nerve disorders, diseases, and basic anatomy. Nerve recovery after transection; nerve compression syndromes; perineurial invasion by tumor; and anatomical concepts are briefly discussed in light of these findings.

### Nerve recovery after transection

*Microsurgical nerve repair does not re-establish distal CSF circulation*. The most devastating injuries are those involving the endoneurium, the terminal CSF drainage to axons. This may explain Wallerian degeneration (degeneration of axons distal to the site of injury). In addition, acute nerve transection may lead to increased intra-neural pressure due to CSF outflow obstruction: a form of *compartment syndrome*.^36^ Proximal nerves exhibit the worst recovery after repair; proximal nerves carry the greatest CSF volume; and proximal nerves are at the greatest risk for intra-neural compartment syndrome after transection. This has enormous implications for spinal cord repair.

### Nerve compression syndromes

The hand surgeon will understand the implications of Figure 2b, where the largest vessels capable of decompressing antegrade CSF flow are found proximal to potential sites of nerve compression (wrist, elbow, neck). An ancillary finding was noted *repeatedly*: *nerves can be compressed by lymphatic networks* not associated with known areas of compression (Figure 12). This may explain residual symptoms after nerve release.

**Figure 12.**
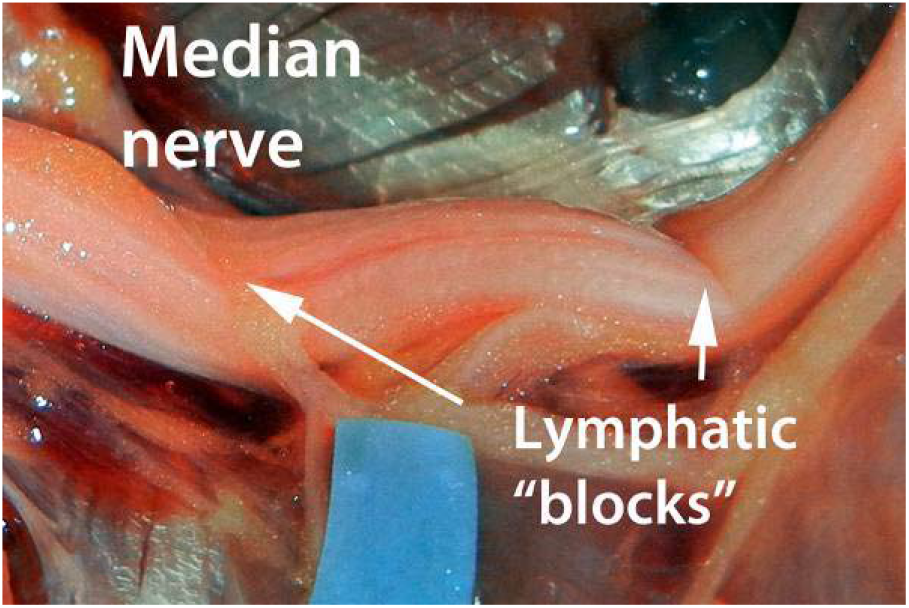
Median nerve compressed solely by a lymphatic plexus.

### Perineurial nerve invasion by tumor (PNI)

Clinicians are familiar with the propensity of *“neurotropic”* tumors (e.g. adenoid cystic carcinoma, melanoma, breast cancer) to invade and migrate proximally along nerves.^8-10^ Research suggests tumor cells invade nerves along a path of least resistance.^8-10,13,14^ This concept reconciles the tissue layers involved but not the direction of tumor propagation (toward the CNS). The Heldermon Lab created a model of PNI in which immortalized breast cancer cells are injected in rat hind limb. Sciatic nerve biopsy confirms tumor cells invade perineurium and migrate proximally. In addition, tumor cells closely *abut and travel along perineurial CSF vessels* (Figure 13). A testable hypothesis is that tumor cells are biologically “attracted” to CSF and migrate proximally towards the highest concentration (i.e. the CNS) of some factor: if this is correct, interruption of CSF flow would result in diminished proximal invasion by tumor cells.

**Figure 13.**
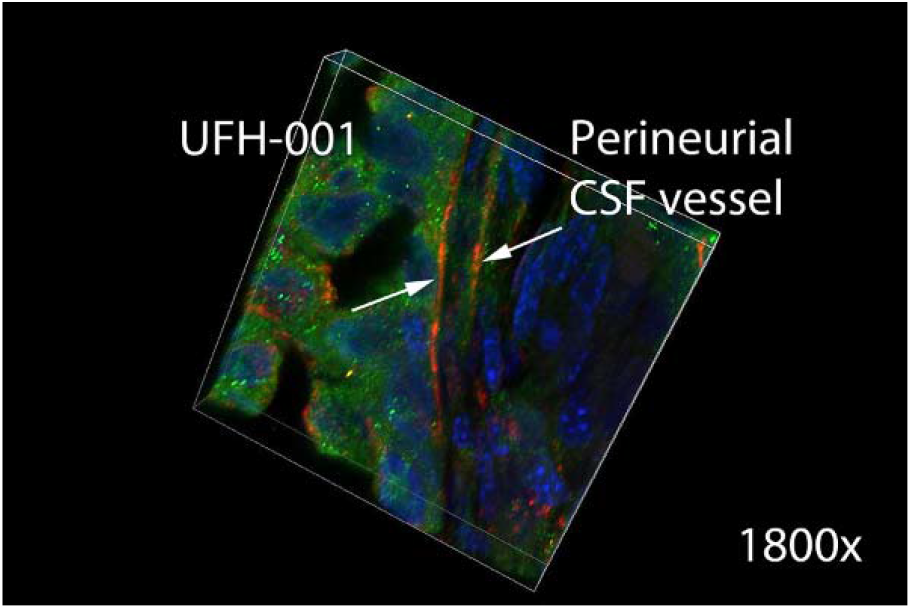
Immortalized breast cancer cells (Luciferase+ green) abut and travel along vimentin+ perineurial CSF channels (red fluorescence indicates vessel walls) in mouse sciatic nerve. 1800x. Volume view assembled from z-stack images 70.10µ x 70.10µ x 10µ; images acquired at 0.2µ intervals, Weber deconvolution.

### Basic anatomical concepts

Two basic anatomical tenets need to be revised and/or amended. (1) Sappey’s Rule should be amended from an *absolute* to a *relative* criterion to characterize a novel vessel as lymphatic.^6^ (2) The current anatomical definition of the carotid sheath should be expanded to include a 4^th^ structure: the terminal CSF drainage of the brain. The proximity between internal jugular (IJ) vein and the CSF drainage of the cerebral hemisphere suggests caution when dissecting the IJ during vascular and microsurgical procedures.

### Sites of periosteal fixation and dural CSF drainage

Loveau et al observed that dural (intra-cranial) CSF vessels drain directly to subcutaneous lymphatics along the sagittal sinus in mice, a novel finding.^32^ Loveau’s work enabled us to identify dural CSF vessels exiting through discrete foramina to drain into subcutaneous lymphatics. Dural CSF vessels (within the cranium) communicate with subcutaneous lymphatics at other sites of periosteal fixation, for example the temporal crest and nuchal ridge. A re-appraisal of the anatomy of periosteal fixation, pioneered by Knize, may be warranted.^37,38^

### The blood-nerve and blood-brain barriers

The blood-nerve and the blood-brain barriers have been the focus of intense research, yet their definitive location and structure has yet to be idenfied.^35,39-41^ Perineurium expresses the insulin-dependent glucose-1 transporter protein (GLUT-1), and has been suggested as the site of blood-nerve interaction.^28^ We can identify GLUT-1 positive CSF vessels in perineurium and arachnoid (Figure 14), *suggesting these vessels as the structural analogue of the blood-nerve and blood-brain barrier*.^42^

**Figure 14.**
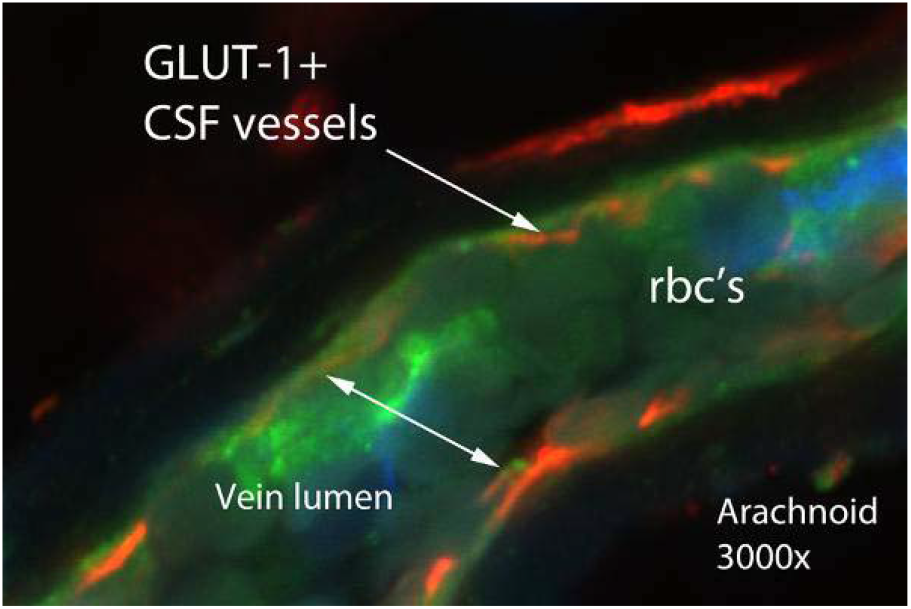
GLUT-1+ CSF vessels surrounding an arachnoid vein in human brain.

### Ancillary findings

Many additional observations were made during the course of this 5-year study. Three of the more relevant are presented as they relate to aesthetic, cleft lip, and plastic surgery.

### The white roll of the lip is determined by a lymphatic vessel

The anatomy of the white roll has been investigated by others.^43^ IR imaging identified a major lymphatic beneath the white roll of the upper lip. This is easily identified by anatomic dissection.

### Lymphatic vessels determine the location of static, dynamic, and senile wrinkles

IR imaging identified lymphatics *beneath* the nasolabial crease, the junction of fat compartments, and rhytides. Additional evidence suggests *all wrinkles are located over lymphatic vessels*, regardless of their designation as static, dynamic, senile and/or involutional.

### Injectables travel further than what is observed using ambient light

Our experience with IR imaging suggests the following: observation of injections using ambient light always underestimates the true extent of diffusion.^44^ The complications of neck weakness, dysphagia, and dysphonia after chemodenervation are directly related to this observation.

### Limitations of this study

Although preliminary flow studies suggest arachnoid CSF travels to endoneurial CSF vessels, this requires additional work. The strength of this study is the anatomy. This work was completed in early 2017, and since then the first author has received additional training in molecular biology. Our ability to image CSF vessels has advanced considerably such that CSF vessels in all layers can now be imaged with greater precision.

Vessels in the human CSF circulatory system are very different from blood vessels and lymphatics. They represent a distinct type of vessel(s). Our observations suggest possible homology with the vascular circulatory system of plants.

### Xylem and phloem

When forced to conclude that we were seeing a novel type of vessel, we searched elsewhere in biology for a potential analogue system; and thought of xylem and phloem. Plants may express F-actin and vimentin exactly like vessels in the human CSF circulatory system.^51-53^ Phloem channels represent a *sucrose-transport* system; CSF vessels a *glucose-transport* system.^54^ Even more intriguing, the circulatory system of plants may express *a biomarker for human blood vessels* (CD105; Figure 15). The authors have additional evidence of biological and structural homology that is outside the scope of this manuscript.

**Figure 15.**
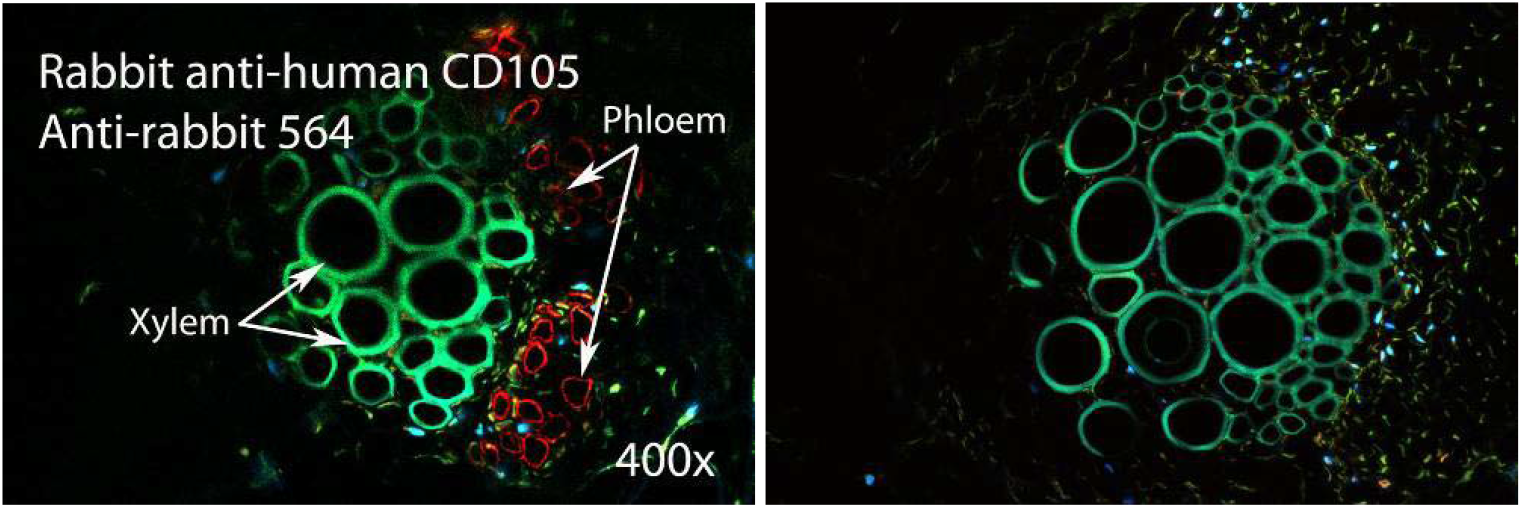
Carota daucus phloem channels test positive against anti-human CD105, a biomarker for human blood vessels. Right, negative control.

Every part of the human CSF circulation will require additional investigation. Since the completion of this body of work in early 2017, it has become clearer that *priangeo vessels* are structurally distinct from blood vessels and lymphatics. Our current work characterizes them as a system cells capable of inter-cellular fluid transfer (Figure 16). Future studies will provide the evidence for this structural concept. This paper was written for the clinical surgeon, and especially for the junior clinical plastic surgeon in hopes of stimulating their interest in this research. The study of the human CSF circulatory system is a field in its infancy, and the well-trained and motivated plastic surgeon can play an important role in its development.

**Figure 16.**
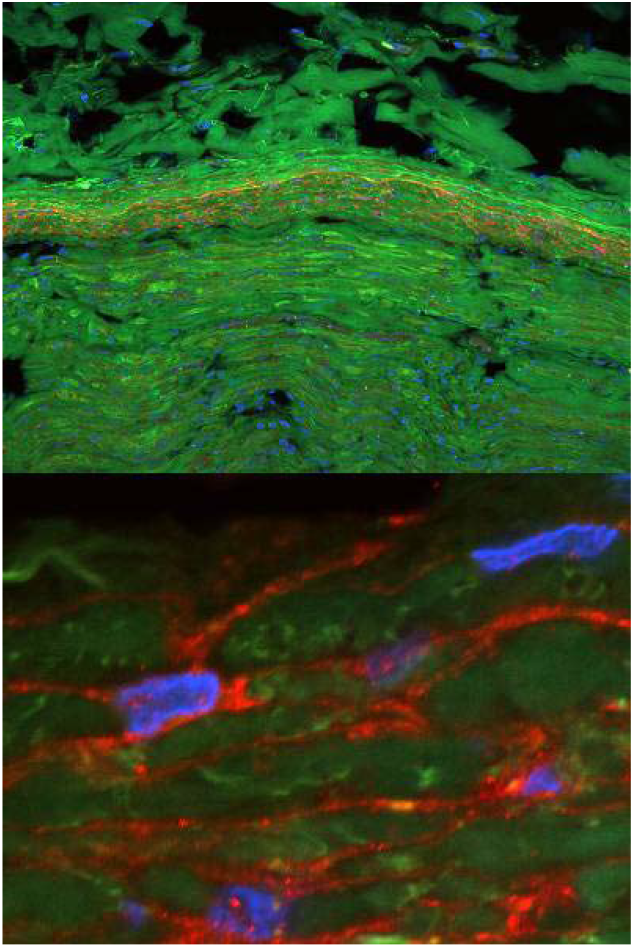
Top. The perineurium expresses the insulin-dependent glucose-1 transporter protein (red fluorescence) and is the likely structural analogue of the physiological blood-nerve barrier. Figure 16, Bottom. At 3000x, the walls of CSF vessels (priangeo vessels) express GLUT-1. Companion cells are located at the junction of multiple vessels. The morphology of CSF vessels suggests that circulatory flow is accomplished by inter-cellular fluid transfer.

## CONCLUSION

**Evidence suggests a novel CSF circulatory system exists in all human nerves**.

## MATERIALS AND METHODS

All antibodies and fluorescent dyes were acquired from Sigma Aldrich, USA unless stated otherwise. AlexaFluor™ secondary antibodies were used (Invitrogen, Carlsbad, CA). Antibodies were verified against human tissues prior to use (e.g. lymphatic antibodies tested against salivary gland). Dilutions were by titration. Positive samples were tested against negative controls (secondary antibody only). Fixation was by acetone, or by paraformaldehyde + antigen retrieval using HIER (citrate buffer). Sectioning followed graded sucrose preservation. All imaging was done using the Nikon AR-1 multi-photon microscope in confocal mode.

### 1. Are epineurial CSF vessels present on all nerves?

Twenty seven (N=54) upper extremities were dissected using the Leica M525 operating microscope (Leica Microsystems, Jena, Germany). The anatomy of epineurial CSF vessels on median and ulnar nerves was evaluated from carpal canal to thoracic outlet.

### 2. Do epineurial CSF vessels have a defined lumen?

Epineurial CSF vessels were identified in the median nerve of 10 upper extremity dissections. Direct intra-luminal injection was performed using various dyes including India ink, indocyanine green, and fluoroscein isothiocyanate (FITC). Results were documented.

### 3. Do epineurial CSF vessels drain into thoracic duct?

A series of 13 hemi-thoracic (extremity/thorax/neck in-continuity) dissections were performed to identify the terminal drainage of CSF from nerves. The brachial plexus was dissected in each specimen, from which CSF communicating vessels traveling from nerve to the thoracic duct were noted. Fluorescent imaging was used to confirm flow to the thoracic duct in selected specimens.

### 4. Do epineurial CSF vessels drain into lymphatics?

Eight upper extremity dissections were prepared by isolating the median nerve at the wrist. A lymphatic vessel was identified (traveling away from the nerve) and injected (retrograde) using both visible dye and infrared fluorescent dye. The ability of 3 raters to identify dye traveling from the lymphatic to an epineural channel on the nerve was evaluated using visible vs. infra-red imaging. Results were compared using Fisher’s Exact Probability Test.

### 5. Do epineurial CSF vessels express a biomarker?

Immunohistochemistry using the known lymphatic biomarkers PROX-1, FOXC2, podoplanin, LYVE-1 failed to define epineurial CSF channels. Vimentin was identified as a candidate biomarker. 1-step direct immunohistochemistry was performed using anti-vimentin antibody (V9 clone conjugated to Cy5) on 28 median nerve specimens that were subsequently imaged with confocal laser microscopy.

### 6. Is vimentin a reliable biomarker for perineurial CSF vessels?

We statistically compared the ability of 3 raters to identify *perineurial* CSF vessels using anti-vimentin antibody compared to the common lymphatic biomarkers LYVE-1 and podoplanin. Eight median nerve specimens were tested using 2-step indirect immunohistochemistry and triplicate hard copy images generated. Statistics were tested with a linear mixed effects model using the F-Test.

### 7. Can we identify the CSF circulation of human brain by anatomic dissection?

We performed gross anatomic dissections in 8 cranial specimens in a non-washout model to preserve filling of blood vessels. Dural and arachnoid CSF vessels were noted.

### 8. What is the terminal CSF drainage from the human brain?

We attempted to define CSF drainage vessels traveling from the skull to the thoracic duct in 13 cadaver dissections (thorax/neck/cranium in-continuity). The right thoracic duct was identified: A CSF vascular plexus (a group of CSF vessels) was found to terminate in the thoracic duct, after which it was dissected cephalad to the cranial base (i.e. known to unknown; retrograde). Anatomical relationships were noted.

### 9. Does CSF drain from the brain to thoracic duct?

This series includes 8 complete dissections (thorax/neck/cranium in continuity). The cervical CSF system was cannulated. Simultaneous craniotomy identified the terminal intra-cranial CSF plexus within the temporal fossa. Visible and infrared dyes were injected from the neck towards the brain. The ability of 3 raters to identify dye traveling from the extra-cranial to the intra-cranial CSF system was evaluated using visible light vs. infra-red imaging. Results were compared using Fisher’s Exact Probability Test.

### 10. What additional biomarkers are expressed by the CSF circulatory system?

We discuss our observations over 5 years using ancillary biomarkers to identify CSF vessels in nerves, within the neck, and in the brain.

## Data Availability

All DATA related to this study is provided. Statistical DATA can be provided upon request.

## ACKNOWLEDGEMENTS

The authors thank the donors and families of the UTSW Willed Body Program. The authors thank the nurses of Zale Lipshy operating room for their help. All molecular imaging was performed at the McKnight Brain Institute. Susan Frost PhD allowed us to use UFH-001 immortalized breast cancer cell line. Special thanks to UF and all the researchers who contributed to this study.

